# Quantifying the Mortality and Morbidity Impact of Medicaid Retractions

**DOI:** 10.1101/2025.05.19.25327564

**Authors:** Abhishek Pandey, Yang Ye, Alison P. Galvani

## Abstract

Recent U.S. legislative proposals include sweeping Medicaid retractions and the expiration of enhanced ACA Premium Tax Credits, threatening health insurance coverage for millions of Americans. Using a validated simulation model, we estimate that 7.7 million individuals becoming uninsured due to the proposed Medicaid changes would lead to a median of 11,308 excess deaths annually. When combined with the 5 million projected to lose coverage due to ACA policy expirations, over 20,000 additional deaths may occur each year. In addition to mortality, coverage loss is projected to result in substantial increases in uncontrolled chronic conditions, including 138,851 additional cases of uncontrolled diabetes, 165,165 cases of uncontrolled hypertension, and 46,200 cases of uncontrolled high cholesterol annually. These projections underscore the wide-reaching public health consequences of limiting access to healthcare.

## Introduction

Recent federal legislative proposals have been introduced to enact sweeping retractions to Medicaid (1). These provisions include delays in the implementation of the Centers for Medicare & Medicaid Services (CMS) Final Rule intended to streamline Medicaid renewals and impose additional administrative requirements for eligibility redeterminations. The Congressional Budget Office (CBO) estimates that the combination of rollbacks outlined in the legislative proposal would result in 7.7 million Americans becoming uninsured (2). In parallel, an additional 5 million individuals are projected to lose insurance due to the scheduled expiration of enhanced Affordable Care Act (ACA) Premium Tax Credits, further compounding the risk of large-scale coverage loss. Although the analysis conducted by the CBO primarily focuses on budgetary implications, far-reaching health consequences of insurance loss include elevated risks of delayed care, unmanaged chronic conditions, and premature mortality (3–5).

In previous work, we developed a simulation-based modeling framework to estimate the excess mortality associated with revisions to policies that affect healthcare coverage, incorporating both current coverage patterns and elevated mortality risk among uninsured individuals.In this study, we extend that framework to estimate the national mortality burden and chronic disease burden associated with the insurance losses resulting from the proposed Medicaid retractions.

## Methods

Building upon our previously established method, we adapted the model to specifically assess the mortality consequences of the anticipated Medicaid coverage reductions(6). While the CBO does not disaggregate the estimate of people losing insurance by age, coverage for children and seniors is largely preserved through other eligibility pathways, whereas the proposed provisions primarily target non-disabled, working-age adults. Therefore, we distributed insurance loss within the 19–64 age group.

We parameterized our simulations with population (7) and insurance coverage (8) data from the United States Census Bureau, adjusting for the estimated loss of insurance for 7.7 million and 13.7 million individuals, respectively, to project annual excess mortality. We first updated insurance coverage within this age group by incorporating the projected loss of coverage, and then applied an elevated mortality hazard ratio (4) associated with being uninsured to estimate the resulting number of annual excess deaths (**Supplementary Appendix**). We incorporated empirical variability in both the hazard ratio and current insurance coverage to generate uncertainty intervals around our projections of excess mortality.

To estimate the morbidity burden, we evaluated three prevalent chronic conditions: diabetes, hypertension, and high cholesterol. For each, we defined control using clinically accepted thresholds: hemoglobin A1c < 8.0% for diabetes, systolic blood pressure < 140 mm Hg for hypertension, and total cholesterol < 240 mg/dL for high cholesterol. Using the same coverage loss projections as in the mortality analysis, we estimated the number of excess cases of uncontrolled disease resulting from insurance loss. For each condition, we applied disease-specific prevalence(9–11) and baseline control rates(9,10,12) to the population losing insurance. We then estimated the number of additional cases of uncontrolled disease attributable to loss of Medicaid coverage by applying estimates of the change in disease control associated with losing health insurance (**Supplementary Appendix)**. For diabetes, we used a relative risk of poor glycemic control among the uninsured compared to insured(13); for hypertension and cholesterol, we used adjusted differences in control rates from national survey data.(5)

## Results

We project that the proposed retractions to Medicaid would cause a median of 11,308 (95% Uncertainty Interval (UI): [2,315, 23,302]) additional deaths annually among individuals aged 19–64 stemming from 7.7 million people becoming uninsured (**Table 1**). Combining those who would lose insurance due to the new legislation and the 5 million who are already scheduled to lose insurance upon expiration of Enhanced ACA Premium Tax Credits (for the total of 13.7 million), we project a median of 20,119 (95% UI: [4,119,41,459]) additional deaths every year.

**Table 1:**
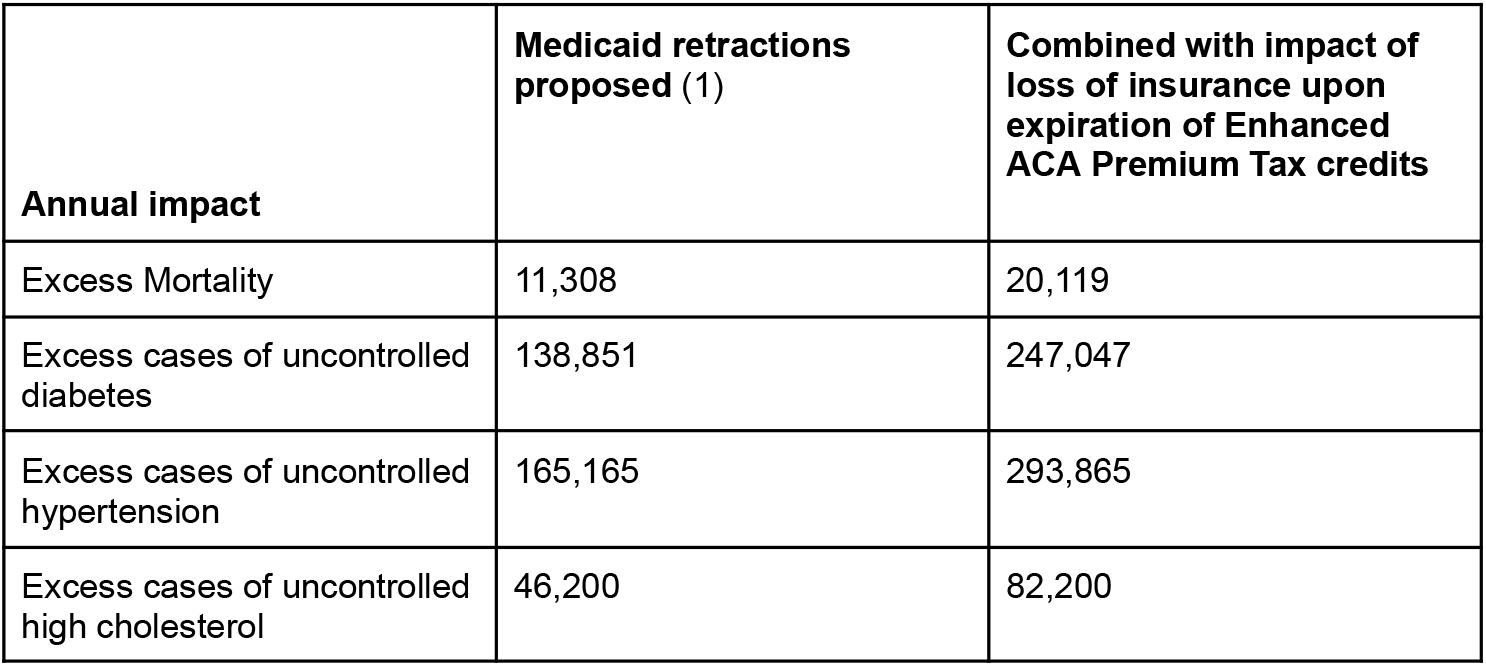
Mortality and morbidity impact of proposed retractions to Medicaid.

Beyond mortality, our model estimates substantial increases in uncontrolled chronic conditions among those who lose insurance (**Table 1**). Under the Medicaid retraction scenario, we project 138,851 additional cases of uncontrolled diabetes, 165,165 additional cases of uncontrolled hypertension, and 46,200 additional cases of uncontrolled high cholesterol annually. When accounting for the additional 5 million losing insurance due to ACA tax credit expiration, the projected morbidity burden rises to 247,047 excess cases of uncontrolled diabetes, 293,865 of uncontrolled hypertension, and 82,200 of uncontrolled high cholesterol.

## Discussion

Medicaid is foundational to the U.S. healthcare system, providing access to essential healthcare services and safeguarding continuity of care for millions of low-income Americans facing treatment barriers. Over the past decade, reforms such as streamlined renewals and expanded eligibility, demonstrably increased insurance coverage, improved chronic disease management, and reduced preventable mortality (3–5). The proposed legislative rollbacks risk undermining these gains. While intended to reduce federal spending, these changes would come at a steep human cost and may, in fact, have counterproductive economic repercussions (14). Our analysis finds that the proposed Medicaid retractions alone could precipitate more than 11,000 additional deaths each year. We also estimate that the scheduled expiration of enhanced ACA Premium Tax Credits would result in more than 8,800 additional deaths annually. Combined, we calculate that over 20,000 unnecessary lives would be lost every year.

In addition to the mortality toll, our estimates indicate that hundreds of thousands more individuals would suffer from uncontrolled chronic conditions. The loss of insurance is projected to result in nearly 140,000 additional cases of uncontrolled diabetes and over 160,000 cases of uncontrolled hypertension annually, with even greater burdens when ACA expirations are included. These conditions substantially increase the risk of heart attack, stroke, renal failure, and other complications, while placing additional strain on the healthcare system.

While our results focus on the mortality implications of losing health insurance, other provisions in the legislation affect mortality through aspects of the bill that even affect people who are insured. For example, Sections 44101–44102 would repeal CMS’ recently finalized rule requiring minimum nursing home staffing. An analysis estimates that enforcing this rule would prevent approximately 12,945 deaths annually by improving staffing and care quality. Blocking its implementation, as proposed in the legislation, would forgo this benefit—representing an additional indirect mortality burden. When combined with our insurance-based estimate of above 20,000 excess deaths annually, the total projected mortality impact could exceed 33,000 preventable deaths per year.

Disrupted coverage jeopardizes disease management, strains emergency care capacity, diminishes workforce productivity, and imposes unsustainable financial burdens on households. Vulnerable individuals, including those facing unstable employment or difficulties navigating complex bureaucratic systems, are likely to bear the brunt of these disruptions. Our findings underscore the insidious public health ramifications of the proposed legislation. Consequently, policymakers must consider not only the fiscal implications of Medicaid reform but also the fundamental public health and equity imperatives at stake. Treating reduced healthcare access as a mere budgetary adjustment disregards the associated human suffering.

## Data Availability

All data produced in the present work are contained in the manuscript.

https://github.com/abhiganit/medicaid-cuts.git

## Reference

1. U.S. Senate Committee on Finance, Majority Staff. [Internet]. [cited 2025 May 13]. Updated memorandum on impacts of proposed Medicaid and Marketplace provisions. Available from: https://d1dth6e84htgma.cloudfront.net/05_13_2025_FCMU_Memorandum_UPDATED_55a74a132a.pdf

2. U.S. House Committee on Energy and Commerce, Democratic Staff. [Internet]. 2025 [cited 2025 May 13]. CBO emails regarding reconciliation scores. Correspondence from the Congressional Budget Office. Available from: https://democrats-energycommerce.house.gov/sites/evo-subsites/democrats-energycommerce.house.gov/files/evo-media-document/cbo-emails-re-e&c-reconcilation-scores-may-11,-2025.pdf#page=1.00

3. Huguet N, Green BB, Larson AE, Moreno L, DeVoe JE. Diabetes and hypertension prevention and control in community health centers: Impact of the Affordable Care Act. J Prim Care Community Health. 2023 Jan;14:21501319231195697.

4. Wilper AP, Woolhandler S, Lasser KE, McCormick D, Bor DH, Himmelstein DU. Health insurance and mortality in US adults. Am J Public Health. 2009 Dec;99(12):2289–95.

5. Hogan DR, Danaei G, Ezzati M, Clarke PM, Jha AK, Salomon JA. Estimating the potential impact of insurance expansion on undiagnosed and uncontrolled chronic conditions. Health Aff (Millwood). 2015 Sep;34(9):1554–62.

6. Pandey A. Github repository. [cited 2025 May 14]. Simulation model to evaluate excess deaths due to loss of insurance coverage. Available from: https://github.com/abhiganit/medicaid-cuts.

7. US Census Bureau. 2023 National Population Projections Datasets. 2023 Nov 9 [cited 2025 Feb 25]; Available from: https://www.census.gov/data/datasets/2023/demo/popproj/2023-popproj.html

8. U.S. Census Bureau. Health insurance coverage status (Table S2701), 2023: ACS 5-year estimates subject tables. American Community Survey [Internet]. [cited 2025 Feb 25]. Available from: https://data.census.gov/table/ACSST5Y2023.S2701?q=health%20insurance

9. CDC. Diabetes. 2024 [cited 2025 May 10]. National Diabetes Statistics Report. Available from: https://www.cdc.gov/diabetes/php/data-research/index.html

10. Fryar CD, Kit B, Carroll MD, Afful J. Hypertension prevalence, awareness, treatment, and control among adults age 18 and older: United States, August 2021-August 2023. NCHS Data Brief [Internet]. 2024 Oct;(511). Available from: https://www.ncbi.nlm.nih.gov/pubmed/40085792

11. Carroll MD, Fryar CD, Gwira JA, Iniguez M. Total and high-density lipoprotein cholesterol in adults: [Internet]. [cited 2025 May 11]. Available from: https://www.cdc.gov/nchs/data/databriefs/db515.pdf

12. Prevalence of Cholesterol Treatment Eligibility and Medication Use Among Adults - United States. 2005.

13. Chasens ER, Dinardo M, Imes CC, Morris JL, Braxter B, Yang K. Citizenship and health insurance status predict glycemic management: NHANES data 2007-2016. Prev Med. 2020 Oct;139(106180):106180.

14. Galvani AP, Parpia AS, Foster EM, Singer BH, Fitzpatrick MC. Improving the prognosis of health care in the USA. Lancet. 2020 Feb 15;395(10223):524–33.

