## Supplementary Appendix for "Quantifying the Mortality and Morbidity Impact of Medicaid Retractions"

### Excess Annual Mortality

Using our previously developed method,(1) we estimated the number of annual excess deaths among Americans who may lose coverage.

If the probability of death for insured individuals in population P in the age-group of interest is μ, then

$$D_c = \mu\, i_c P + \lambda \mu (1 - i_c) P,$$

where $i_c$ is the current insured proportion within this age group and λ is the hazard ratio of death among uninsured individuals compared to insured individuals. We used data from the United States Census Bureau to inform the current insured proportion ($i_c$)(2) .

If n is the number of Medicaid beneficiaries in population P who would become uninsured as a result of policy change, the insurance coverage among individuals in population P would change. We calculate the updated insurance coverage as:

$$i_n = i_c - \frac{n}{P},$$

The expected number of annual deaths becomes:

$$D_n = \mu\, i_n P + \lambda \mu (1 - i_n) P,$$

or equivalently:

$$D_n = \frac{i_n + \lambda\ (1 - i_n)}{i_c + \lambda\ (1 - i_c)} D_c$$

The additional annual deaths are then calculated as $D_n - D_c$ .

### Excess Annual Uncontrolled Morbidity

Using data on disease prevalence, clinical control rates, and the observed effect of insurance coverage on disease management, we project changes in population-level health burdens as a result of change in insurance coverage. We defined disease control status using clinically accepted thresholds in alignment with federal health datasets. Specifically, diabetes was considered controlled if individuals with a diabetes diagnosis had hemoglobin A1c levels below 8.0%. Hypertension control was defined by systolic blood pressure below 140 mm Hg among those diagnosed with high blood pressure. For high cholesterol, control was defined as total cholesterol below 240 mg/dL among individuals with a diagnosis of high cholesterol. Control rates represent the share of diagnosed individuals whose condition is considered clinically managed, while the uncontrolled rate reflects the remainder who do not meet these thresholds.

Let P denote the size of the population under study, and let p represent the prevalence of a given chronic disease in that population. Let $i_c$ and $i_n$ represent the insurance coverage rates before and after a policy change respectively. We define $r_i$ as the proportion of insured individuals with uncontrolled disease, and $r_u$ as the corresponding proportion among uninsured individuals.

The total number of uncontrolled cases prior to coverage loss is:

$$U_{current} = p\,P\,(i_c r_i + (1 - i_c) r_u)$$

After the policy-induced change in insurance coverage, the total number of uncontrolled cases becomes:

$$U_{updated} = p\,P\,(i_n r_i + (1 - i_n) r_u)$$

The excess uncontrolled burden resulting from the change is then:

$$\Delta U = U_{updated} - U_{current} = p\,P\,(i_c - i_n)\,(r_u - r_i)$$

This formulation provides a general framework for estimating excess cases of uncontrolled disease as a result of policies that reduce insurance coverage. To populate this model, we drew prevalence and control data from recent national datasets stratified by age group.(3) (4) (5)
In the absence of direct data on the proportion of adults with controlled high cholesterol, we used the proportion of individuals with high cholesterol who reported taking cholesterol-lowering medication(6)

To estimate the shift in control associated with insurance loss, we used two approaches depending on data availability. For diabetes, we applied a relative risk-based framework. We first converted an adjusted odds ratio of 1.99 for poor glycemic control among uninsured adults compared to insured adults(7) to a relative risk using the formula:

$$R = \frac{OR}{1-p_0 + p_0 OR},$$

where $p_0 = 0.074$ is the proportion of insured adults with uncontrolled diabetes in the same study. This conversion yielded a relative risk of 1.86. The rate of uncontrolled diabetes among the uninsured is then calculated as: $r_u = R\, r_i$.

Substituting this into the expression for the number of individuals with uncontrolled diabetes, we obtain:

$$U_{updated} = \frac{i_n + R(1-i_n)}{i_c + R(1-i_c)} U_{current}$$

where:

$$U_{current} = p\, P\, r,$$

with p as disease prevalence, P as population size, and r as the overall uncontrolled rate of diabetes in the population. The difference ($\Delta U$) between $U_{updated}$ and $U_{current}$ reflects the excess number of uncontrolled cases attributable to insurance loss.

For hypertension and high cholesterol, we used adjusted differences in control rates between insured and uninsured adults, derived from nationally representative survey data(8). The rate of uncontrolled disease among uninsured individuals is expressed as:

$$r_u = r_i + \Delta,$$

where Δ represents the adjusted difference in disease control (0.055 for hypertension, 0.050 for cholesterol) between the uninsured and insured. These differences were applied directly to estimate the increase in uncontrolled disease attributable to the coverage reduction.

Together, this approach quantifies the policy-induced health burden in terms of excess unmanaged chronic conditions, offering a complementary lens to mortality-based impact estimates.

Reference:


1. Pandey A, Fitzpatrick MC, Singer BH, Galvani AP. Mortality and morbidity ramifications of proposed retractions in healthcare coverage for the United States. Proc Natl Acad Sci U S A. 2024 Apr 30;121(18):e2321494121.

2. U.S. Census Bureau. Health insurance coverage status (Table S2701), 2023: ACS 5-year estimates subject tables. American Community Survey [Internet]. [cited 2025 Feb 25]. Available from: https://data.census.gov/table/ACSST5Y2023.S2701?q=health%20insurance

3. CDC. Diabetes. 2024 [cited 2025 May 10]. National Diabetes Statistics Report. Available from: https://www.cdc.gov/diabetes/php/data-research/index.html

4. Fryar CD, Kit B, Carroll MD, Afful J. Hypertension Prevalence, Awareness, Treatment, and Control Among Adults Age 18 and Older: United States, August 2021–August 2023 [Internet]. National Center for Health Statistics (U.S.); 2024 Dec. Available from: https://www.cdc.gov/nchs/data/databriefs/db511.pdf

5. Carroll MD, Fryar CD, Gwira JA, Iniguez M. Total and high-density lipoprotein cholesterol in adults: [Internet]. [cited 2025 May 11]. Available from: https://www.cdc.gov/nchs/data/databriefs/db515.pdf

6. Prevalence of Cholesterol Treatment Eligibility and Medication Use Among Adults — United States, 2005–2012 [Internet]. 2015 [cited 2025 May 11]. Available from: https://www.cdc.gov/mmwr/preview/mmwrhtml/mm6447a1.htm#tab1

7. Chasens ER, Dinardo M, Imes CC, Morris JL, Braxter B, Yang K. Citizenship and health insurance status predict glycemic management: NHANES data 2007-2016. Prev Med. 2020 Oct;139(106180):106180.

8. Hogan DR, Danaei G, Ezzati M, Clarke PM, Jha AK, Salomon JA. Estimating the potential impact of insurance expansion on undiagnosed and uncontrolled chronic conditions. Health Aff (Millwood). 2015 Sep;34(9):1554–62.
